# Using Real World Data to Understand HIV and COVID-19 in the U.S.A. and Spain: Characterizing Co-Infected Patients Across the Care Cascade

**DOI:** 10.1101/2020.11.10.20229401

**Authors:** Julianna Kohler, Kristin Kostka, Rupa Makadia, Roger Paredes, Talita Duarte-Salles, Scott Duvall, Alison Cheng, Asieh Golozar, Jennifer C. E. Lane, Anthony G. Sena, Peter R. Rijnbeek, Daniel R. Morales, Patrick B. Ryan, Christian Reich, Michael E. Matheny, Kristine E. Lynch, George K. Siberry, Daniel Prieto-Alhambra

## Abstract

**Objective:** Most patients severely affected by COVID-19 have been elderly and patients with underlying chronic disease such as diabetes, cardiovascular disease, or respiratory disease. People living with HIV (PLHIV) may have greater risk of contracting or developing severe COVID-19 due to the underlying HIV infection or higher prevalence of comorbidities.

**Design:** This is a cohort study, including PLHIV diagnosed, hospitalized, or requiring intensive services for COVID-19.

**Methods:** Data sources include routine electronic medical record or claims data from the U.S. and Spain. Patient demographics, comorbidities, and medication history are described.

**Results:** Four data sources had a population of HIV/COVID-19 coinfected patients ranging from 288 to 4606 lives. PLHIV diagnosed with COVID-19 were younger than HIV-negative patients diagnosed with COVID-19. PLHIV diagnosed with COVID-19 diagnosis had similar comorbidities as HIV-negative COVID-19 patients with higher prevalence of those comorbidities and history of severe disease. Treatment regimens were similar between PLHIV diagnosed with COVID-19 or PLHIV requiring intensive services.

**Conclusions:** Our study uses routine practice data to explore HIV impact on COVID-19, providing insight into patient history prior to COVID-19. We found that HIV and COVID-19 coinfected patients have higher prevalence of underlying comorbidities such as cardiovascular and respiratory disease as compared to HIV-negative COVID-19 infected patients. We also found that, across the care cascade, co-infected patients who received intensive services were more likely to have more serious underlying disease or a history of more serious events as compared to PLHIV who were diagnosed with COVID-19.

## Background

The first case of novel coronavirus disease 2019 (COVID-19), caused by the severe acute respiratory syndrome coronavirus 2 (SARS-CoV-2), was detected in Wuhan, China in December of 2019^i^. COVID-19 has since spread across the world, causing nearly 38 million infections and 1.1 million deaths^ii^. Most patients affected by this disease are elderly and/or have underlying chronic diseases such as cardiovascular disease, diabetes, or respiratory disease^iii,iv,v,vi^.

While HIV is frequently listed as a potential risk factor for sever COVID-19 outcomes due to the damage it wreaks on the immune system, it is not clear whether people living with HIV (PLHIV) are at increased risk of COVID-19. PLHIV, even when HIV is controlled with antiretroviral therapy (ART), are at higher risk of many of the conditions implicated in greater risk of more severe COVID-19, such as cardiovascular disease, diabetes, and hypertension.^vii^,^viii^,^ix^ In addition, 19% of PLHIV worldwide do not know their status and, therefore, have uncontrolled disease which may increase their risk of more severe COVID-19^x^.

A study in the Western Cape province of South Africa found that PLHIV had a higher risk of hospitalization and death with COVID-19 than those without HIV^xi^. However, studies from the United States (U.S.), the United Kingdom (U.K.), and Spain found that HIV was not a risk factor for severe COVID-19 outcomes^xii^,^xiii^,^xiv^,^xv^. More recent research has looked at larger populations of PLHIV, generally showing that PLHIV have higher inflammatory markers and may have slightly higher rates of severe outcomes, although these rates tend not to be statistically significant^xvi^,^xvii^,^xviii^. Building on these findings, the purpose of this study was to characterize PLHIV who are co-infected with COVID-19 using real world data from multiple sources in the U.S. and Spain. Specifically, this study explores:

- How are PLHIV who get COVID-19 different than HIV-negative patients who get COVID-19?
- How are PLHIV with COVID-19 (inclusive of all care levels) different than PLHIV with COVID-19 who require intensive services (critical COVID-19)?
- How are PLHIV with critical COVID-19 (hospitalization requiring intensive services) different than HIV-negative patients with critical COVID-19?
- Are there differences in HIV drug use between PLHIV with COVID-19 and PLHIV with critical COVID-19?

## Methods

### Study Design

This study used routinely collected inpatient and outpatient health data collected from January 2020 to September 2020 in the U.S. and Spain. This study was part of CHARYBDIS (Characterizing Health Associated Risks, and Your Baseline Disease In SARS-COV-2), a broader observational cohort study characterizing COVID-19 outcomes and patient history across multiple subgroups. This study was executed by the Observational Health Data Sciences and Informatics (OHDSI) community, a multi-stakeholder, interdisciplinary collaborative to bring out the value of health data through large-scale analytics^xix^. OHDSI maintains the Observational Medical Outcomes Partnership (OMOP) Common Data Model (CDM) and associated tools. A common data model enables semantic interoperability across data sources, allowing analyses to be consistent across data sources. The result set for these analyses was extracted on September 30, 2020.

All data have been mapped to the OMOP CDM. The analytical code developed and utilized in the study is publicly available (https://github.com/ohdsi-studies/Covid19CharacterizationCharybdis).

### Study Population

As part of the CHARYBDIS study, patients were identified as having COVID-19 if they had a record of a positive test for SARS-CoV-2 after December 1, 2019 or had a record of a COVID-19 diagnosis (index date is date of the first of these two events). Patients were defined as hospitalized with a COVID-19 diagnosis if they had any inpatient stay or emergency room visit with an inpatient stay and a positive test for SARS-CoV-2 or a record of COVID-19 diagnoses between 21 days before and up to the final day of the inpatient stay. Patients were defined as hospitalized and requiring intensive services if they had a record of any of 46 services (please see Appendix A for full list), including such procedures as tracheostomy, intubation, respiratory ventilation, or ECMO, an inpatient stay and a positive test for SARS-CoV-2 or a record of COVID-19 diagnoses between 21 days before and up to the final day of the inpatient stay. Note that the COVID-19 diagnosis group is inclusive of the hospitalized and the intensive services group.

In order to be included in the study, patients had to have at least one continuous year of data (continuous enrollment for claims data or an equivalent observation period for electronic medical record data) included prior to the inpatient stay and COVID-19 diagnosis.

Within each cohort (COVID-19 diagnosis, diagnosis with hospitalization, and diagnosis with hospitalization and intensive services), the patient history was scanned for evidence of HIV infection and treatment. The group of PLHIV included individuals with at least one occurrence of an HIV diagnosis (excluding HIV-2). In the U.S. databases, PLHIV were further defined as having at least one occurrence of an HIV drug in addition to the diagnosis.

#### Sample size calculation

A minimum of 140 subjects was deemed necessary a priori to estimate with sufficient precision (confidence interval width of +/- 5%) the prevalence of a previous condition affecting 10% of the study population. Any data source/s or stratum with fewer than 140 subjects was therefore excluded and is not reported as part of this analysis. Of 18 databases included in CHARYBDIS, four had sufficient PLHIV for inclusion in this study: From the U.S., Health Verity (n=457)^xx^, the U.S. Veterans Administration (n=699)^xxi^, IQVIA Open Claims (n=4606)^xxii^, and the Catalonia-based SIDIAP (n=288)^xxiii^. Only IQVIA Open Claims had sufficient data to characterize patient history for co-infected patients who required hospitalization and intensive care services.

### Patient History

To explore prevalent comorbidities in PLHIV with COVID-19, we used disease definition phenotypes developed for the CHARYBDIS study across a variety of conditions and searched for evidence of those phenotypes in the 365 days prior to index date (for more information on phenotypes, visit https://data.ohdsi.org/Covid19CharacterizationCharybdis/). Of interest were comorbidities already established as risk factors for severe COVID-19 infection, such as cardiovascular disease, respiratory disease, diabetes, and kidney disease. We also searched drug exposures in the 365 days prior to the index date to compare the frequencies of HIV treatments in patients who required intensive services to frequencies in all co-infected patients. Of particular interest were tenofovir, emtricitabine, and protease inhibitors, based on evidence of their potential benefit in people with SARS or COVID-19^xxiv^,^xxv^,^xxvi^.

## Results

### How are PLHIV with COVID-19 different than HIV-negative patients with COVID-19?

Within the four included data sources, COVID-19 impacted a varying proportion of PLHIV, from a low of 0.34% of PLHIV being diagnosed with COVID-19 in the IQVIA Open Claims dataset to a high of 5.4% of PLHIV being diagnosed with COVID-19 in the HealthVerity database.

Consistent with the HIV epidemics in both countries, approximately 55-60% of PLHIV with COVID-19 were 50-69 years old (within the SIDIAP database, 63% of patients were 40-59 years old, with only 8% of patients being 60-69 years old). HIV/COVID-19 co-infection was exceedingly rare in children under 20 years old, and co-infected patients over 80 years old were also unusual. HIV-negative COVID-19 patients were generally older than PLHIV co-infected with COVID-19. Co-infected patients were more likely to be male than female (see Table 1).

**Table 1.** Baseline Characteristics of study participants, stratified by data source and HIV status

| Characterization | Data Source |  |  |  |  |  |  |  |
| --- | --- | --- | --- | --- | --- | --- | --- | --- |
|  | Health Verity PLHIV<br>(n=457) | HealthVerity<br>non-HIV<br>(n=113, 716) | VA-OMOP PLHIV<br>(n=699) | VA-OMOP non-HIV<br>(n=54,858) | IQVIA Open<br>Claims PLHIV<br>(n=4 606) | IQVIA Open<br>Claims non-HIV<br>(n=461,585) | SIDIAP (Spain)<br>PLHIV<br>(n=288) | SIDIAP (Spain)<br>non-HIV<br>(n=121,853) |
| Total Patient Lives<br>(non-COVID-19 & COVID-19) | 8,421 |  | 54,150 |  | 1,349,258 |  | 15,134 |  |
| age group: 00-04 |  | 0.2% |  |  | <0.1 | 0.8% |  | 1.2% |
| age group: 05-09 |  | 0.5% |  | 0.0% | 0.2% | 0.7% |  | 1.1% |
| age group: 10-14 |  | 0.8% |  | 0.0% | <0.1 | 0.8% |  | 0.9% |
| age group: 15-19 |  | 1.7% |  | 0.0% | 0.2% | 1.4% |  | 1.3% |
| age group: 20-24 | <1.1% | 4.4% |  | 0.6% | 0.8% | 3.1% | <1.7% | 3.6% |
| age group: 25-29 | 3.1% | 6.4% | 1.4% | 3.2% | 3.2% | 4.5% | 4.2% | 5.4% |
| age group: 30-34 | 5.7% | 7.2% | 3.6% | 6.3% | 4.8% | 5.5% | 7.3% | 6.3% |
| age group: 35-39 | 4.8% | 7.8% | 4.7% | 7.3% | 5.5% | 5.9% | 12.2% | 8.4% |
| age group: 40-44 | 6.8% | 8.1% | 3.6% | 6.4% | 6.4% | 6.3% | 12.8% | 10.7% |
| age group: 45-49 | 9.2% | 9.2% | 6.4% | 6.8% | 8.6% | 7.1% | 18.1% | 10.5% |
| age group: 50-54 | 15.3% | 11.3% | 12.6% | 9.3% | 12.7% | 8.5% | 17.4% | 9.5% |
| age group: 55-59 | 18.8% | 12.1% | 16.9% | 9.8% | 16.1% | 9.6% | 14.9% | 8.6% |
| age group: 60-64 | 18.8% | 11.1% | 17.5% | 11.2% | 15.0% | 9.6% | 4.5% | 6.8% |
| age group: 65-69 | 10.7% | 7.5% | 13.9% | 9.9% | 10.5% | 8.2% | 3.5% | 4.2% |
| age group: 70-74 | 4.2% | 4.7% | 13.2% | 14.1% | 7.6% | 7.2% | 2.1% | 4.0% |
| age group: 75-79 | <1.1% | 3.1% | 4.3% | 7.0% | 4.1% | 6.1% | <1.7% | 4.0% |
| age group: 80-84 | <1.1% | 2.2% | 1.1% | 3.2% | 2.1% | 5.2% | <1.7% | 3.9% |
| age group: 85-89 |  |  | <0.7% | 2.8% | 1.9% | 9.6% | <1.7% | 4.7% |
| age group: 90+ |  | 1.8% | <0.7% | 2.2% |  |  | <1.7% | 4.9% |
| gender = female | 23.6% | 56.3% | 3.9% | 18.5% | 34.7% | 54.9% | 29.5% | 57.9% |
| gender = male | 76.4% | 43.7% | 96.1% | 81.5% | 65.3% | 45.0% | 70.5% | 42.1% |
| Hospitalized | 9.2% | 5.7% | 23.6% | 16.6% | 37.8% | 27.8% | 15.6% | 13.2% |
| Intensive Services | <1.1% | 0.3% | 4.0% | 2.8% | 5.6% | 3.9% | Not available | Not available |

The databases also showed disparities in the proportion of patients hospitalized with a COVID-19 diagnosis (hospitalization could happen anywhere from 0-30 days after COVID-19 diagnosis, so it is not considered an outcome). Across all databases, a higher proportion of PLHIV than HIV-negative patients were hospitalized for COVID-19. The proportion of PLHIV with COVID-19 who required intensive services was similar to HIV-negative COVID-19 patients who required intensive services, although there were only small numbers of patients in this category across datasets. The IQVIA Open Claims dataset was consistent with these trends: 37.8% of PLHIV with COVID-19 were hospitalized, whereas 27.8% of HIV-negative patients with COVID-19 were hospitalized; 5.6% of PLHIV with COVID-19 required intensive services, whereas 3.9% of HIV-negative patients with COVID-19 required intensive services (see Table 1).

### How are PLHIV with COVID-19 (inclusive of all care levels) different than PLHIV with COVID-19 who require intensive services (critical COVID-19)?

Within the IQVIA Open Claims database, we compared the medical history of PLHIV requiring intensive services, as compared to PLHIV infected with COVID-19 (including all levels of care). The most common comorbidities in the intensive services group were largely the same for PLHIV as they are for the HIV-negative population of patients with COVID-19: heart disease, respiratory conditions, diabetes and kidney disease were all more prevalent in patients who received intensive services than in the broader population of patients who were diagnosed with COVID-19 (see Figure 1).

**Figure 1.**
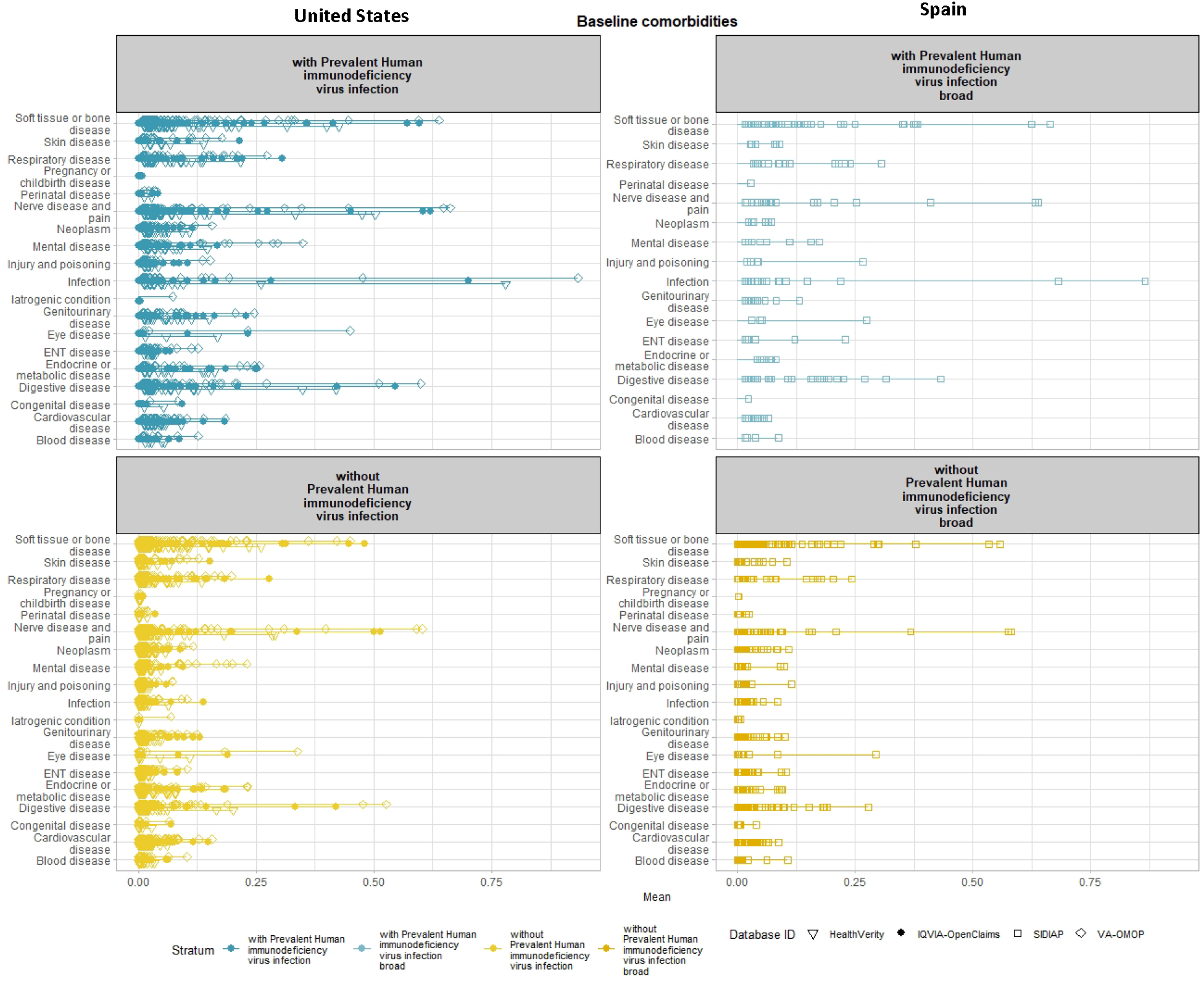
Comparison of baseline comorbidities of PLHIV with a diagnosis of COVID-19 or test positive for SARS-CoV-2

Prevalent comorbidities and history of acute events prior to COVID-19 diagnosis were more frequent in PLHIV with critical COVID-19 than in PLHIV with COVID-19. A history (pre-COVID-19) of cardiovascular events (including acute myocardial infarction, sudden cardiac death, ischemic stroke, intracranial bleed hemorrhagic stroke, heart failure and hospitalization) was present in 10% of PLHIV with COVID-19 but in 21% of co-infected patients who received intensive services. A history of acute myocardial infection itself was present in only 2% of PLHIV with COVID-19 but 8% of co-infected critical COVID-19 patients (see Figure 2).

**Figure 2.**
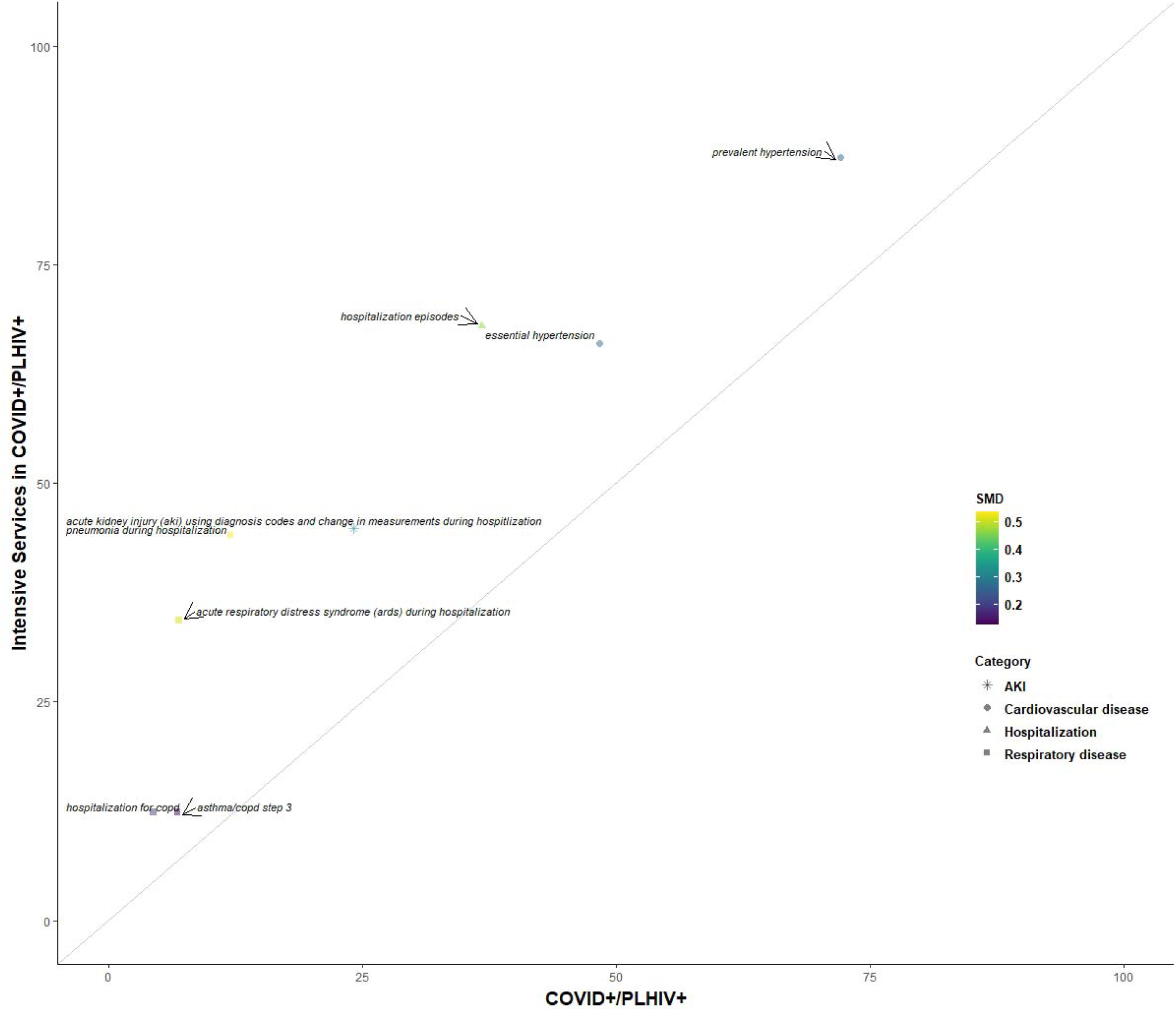
Comparison of baseline characteristics of PLHIV receiving intensive services with a diagnosis of COVID-19 or test positive for SARS-CoV-2 vs. PLHIV with a diagnosis of COVID-19 or test positive for SARS-CoV-2

More severe respiratory disease tends to be more prevalent in PLHIV with critical COVID-19, as compared to PLHIV with COVID-19. For example, mild asthma/COPD (requiring short-acting muscarinic antagonist or short-acting β-agonist treatment or a combination of the two) is no more prevalent in PLHIV with COVID-19 than those requiring intensive services (15.4% vs. 16.1%). But for severe asthma/COPD, requiring long-acting muscarinic antagonist + long-acting β-agonist treatment + inhaled corticosteroid treatment, the prevalence among all co-infected patients is 6.8% versus 12% of patients requiring intensive services. Similarly, 4% of PLHIV with COVID-19 have a history of hospitalization for COPD versus 12% among co-infected patients requiring intensive services. This appears consistent with HIV-negative patients (see Figure 2).

A pre-COVID-19 history of acute kidney disease has the most dramatic differences in prevalence across the disease progression cascade. Only 7% of PLHIV with a COVID-19 diagnosis have AKI, and that prevalence rises to nearly 21% for co-infected patients requiring intensive services; when AKI is identified through laboratory values, the prevalence is 24% and 45% for COVID-19 diagnosed and intensive services PLHIV, respectively. Prevalent chronic kidney disease is present in 23% and 44% of co-infected and intensive services patients, respectively (see Figure 2).

### How are PLHIV with critical COVID-19 (hospitalization requiring intensive services) different than HIV-negative patients with critical COVID-19?

A history of more severe cardiovascular events was more prevalent in PLHIV with critical COVID as compared to HIV-negative patients requiring intensive services: only 1.4% of PLHIV diagnosed with COVID-19 and requiring intensive services had a history of AMI, and 4.5% of HIV-negative patients requiring intensive services. Similarly, 3.5% of co-infected patients had a history of deep vein thrombosis (versus 1.9% of HIV-negative COVID-19 patients), and 7.3% of PLHIV with critical COVID-19 had a history of DVT, compared to 3.7% of HIV-negative patients requiring intensive services. Hypertension and heart disease, however, were more similar across both groups (55% versus 45.8% of non-critical COVID-19 patients and 66.1% versus 66.4% of intensive services patients for heart disease; 72.1% versus 59% of non-critical COVID-19 patients and 87% versus 81.8% of critical COVID-19 patients for hypertension) (see Figure 3).

**Figure 3.**
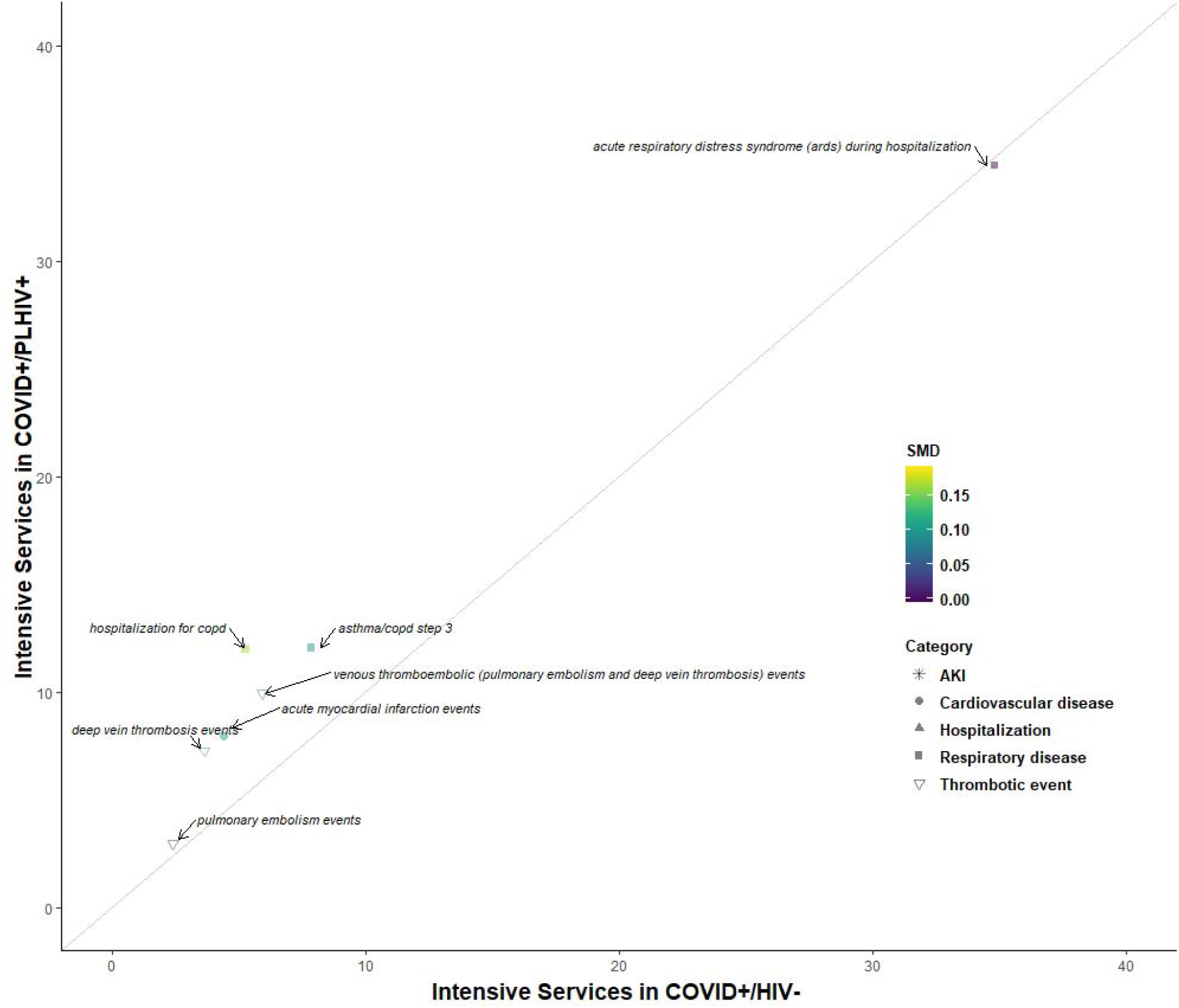
Comparison of baseline characteristics of PLHIV receiving intensive services with a diagnosis of COVID-19 or test positive for SARS-CoV-2 vs. HIV-negative patients hospitalized receiving intensive services with a diagnosis of COVID-19 or a test positive for SARS-CoV-2

### Are there differences in HIV drug use between PLHIV with COVID-19 and PLHIV with critical COVID-19?

For drug history, PLHIV with a COVID-19 diagnosis and PLHIV with COVID-19 and requiring intensive services were generally similar. There were few antiretroviral therapy drugs that were differentially represented across the two groups: cobicistat (a booster) was the most dramatic, at 11.5% of intensive services patients having an exposure in the last year to cobicistat and 13.8% of diagnosed patients having an exposure. Similarly, 7.3% of intensive services patients had an exposure to tenofovir disoproxil in the last year, compared to 8.6% of diagnosed patients. Given the small sample sizes, these differences likely to amount to two to three patients in each group (see Figure 4).

**Figure 4.**
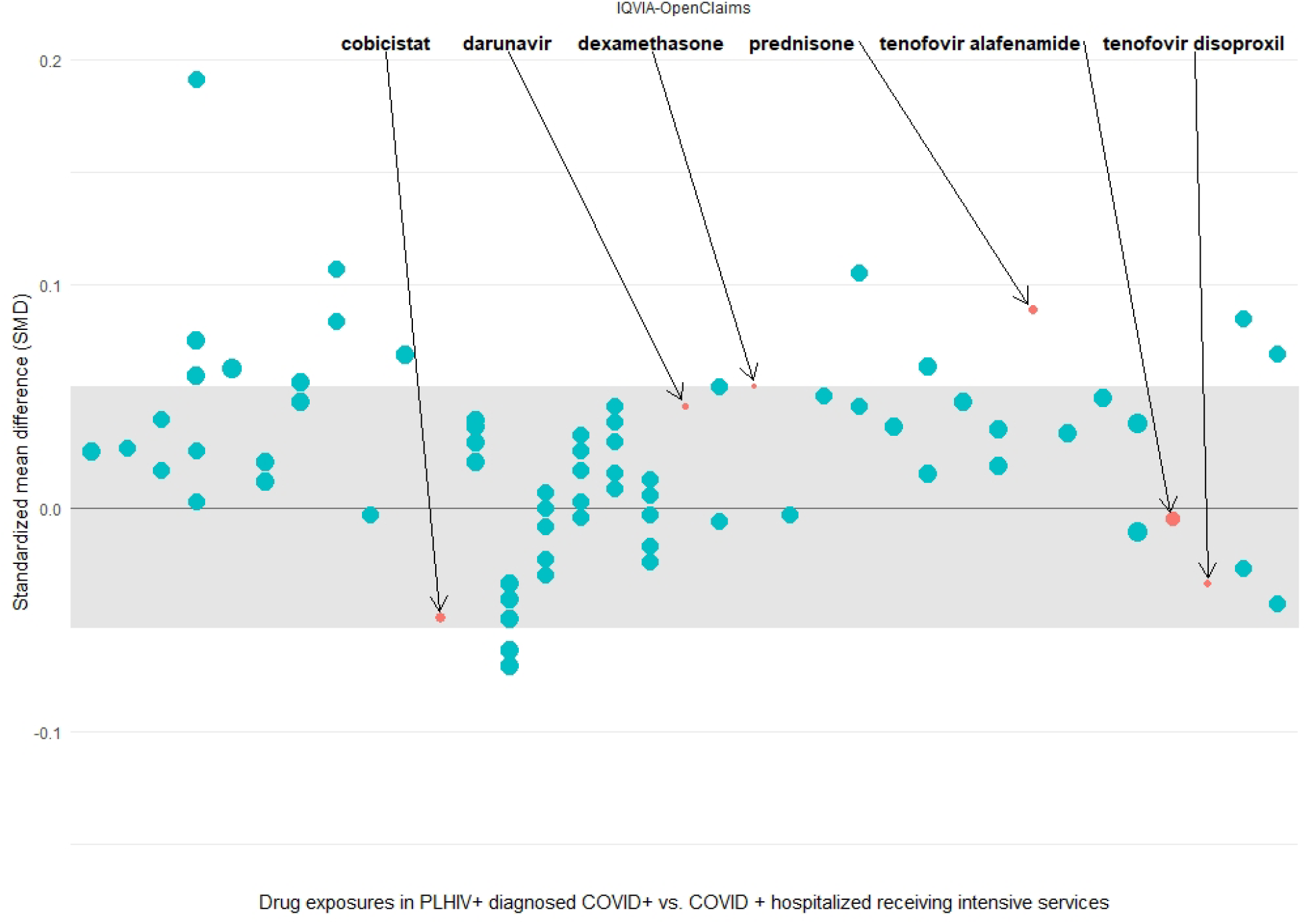
Comparison of baseline treatments of PLHIV receiving intensive services with a diagnosis of COVID-19 or test positive for SARS-CoV-2 vs. PLHIV with a diagnosis of COVID-19 or test positive for SARS-CoV-2

## Discussion

This study characterizes people living with HIV who are diagnosed with COVID-19, including those who are hospitalized and require intensive services. This study found that the comorbidities most prevalent in HIV/COVID-19 co-infected patients are many of the same comorbidities that impact HIV-negative COVID-19 patients, particularly cardiovascular disease, respiratory disease, diabetes, and kidney disease. In addition, more severe comorbidities, or a history of more acute events, such as acute myocardial infarction, hospitalization for COPD, and acute kidney infection, are more prevalent in HIV/COVID-19 co-infected patients than in HIV-negative patients and are also more prevalent in PLHIV with severe COVID-19 (requiring intensive services) than in PLHIV diagnosed with COVID-19, only. This may be caused by a variety of factors, including population size, the known increased risk of chronic diseases associated with HIV, the care patterns that govern HIV treatment, or other factors.

In addition, we found that treatment in the year prior to COVID-19 diagnosis was not markedly different between PLHIV who required intensive services and PLHIV who were only diagnosed with COVID-19, which implies that the regimen may not prevent more severe disease in PLHIV. We also explored drug history to see if there was any evidence that advanced HIV disease is different across COVID-19 severity, as evidenced by use of atovaquone, dapsone, or cotrimoxazole, but we did not see significant use of these drugs across any cohorts.

This study is not designed to explore the impact of HIV on the natural history of COVID-19 disease, but it provides some directional areas for exploration. Overall, PLHIV with COVID-19 are younger than HIV-negative COVID-19 patients; that may be driven by the nature of the HIV epidemic (in most countries, the age distribution of PLHIV is concentrated among adults and less concentrated among older populations and children, as compared to the HIV-negative population), or it may be that HIV confers some level of additional risk to people in middle age. The proportion of HIV-positive patients who are diagnosed with COVID-19 across each of the data sets seems likely to reflect the COVID-19 burden in those areas (for example, nearly 14% of PLHIV were diagnosed with COVID-19 in the Catalonia-based data source, SIDIAP, which is consistent with the high rates of infection observed in Catalonia) rather than differences in the nature of HIV infection in Catalan versus the U.S.

## Conclusion

The HIV community has been concerned since the beginning of COVID-19 pandemic about the implication of HIV treatment on COVID-19 patients, but there has been limited research to date about the impact of HIV. This study characterizes HIV and COVID-19 infected patients across two countries and provides a more in-depth characterization of co-infected patients as they move through the care cascade.

This study found that a higher proportion of PLHIV with common comorbid risk factors for COVID such as diabetes, cardiovascular disease, respiratory disease, and kidney disease ended up with severe disease requiring hospitalization or intensive services, as compared to HIV-negative patients with the same comorbid conditions. More research should explore these factors to understand if they are driven by the nature of the epidemic—PLHIV are at higher risk of these comorbid conditions—or if HIV combined with these comorbidities incurs excess risk to PLHIV.

## Data Availability

Analyses were performed locally in compliance with all applicable data privacy laws. Although the
underlying data is not readily available to be shared, authors contributing to this paper have direct access to the data sources used in this study. All results (e.g. aggregate statistics, not presented at a patient-level with redactions for minimum cell count) are available for public inquiry. These results are inclusive of site-identifiers by contributing data sources to enable interrogation of each contributing site. All analytic code and result sets are made available at: https://github.com/ohdsistudies/Covid19CharacterizationCharybdis

## Acknowledgements

We thank the entire Observational Health and Data Sciences Community, specifically contributions from collaborators: Paula Casajust, Martina Recalde, Elena Roel, Carlos Areia, and Thamir M. Alshammari who provided editorial support to the final manuscript. We would like to acknowledge the patients who suffered from or died of this devastating disease and their families and caregivers. We would also like to thank the healthcare professionals involved in the management of COVID-19 during these challenging times, from primary care to intensive care units.

## Ethical approval

All the data partners received Institutional Review Board (IRB) approval or exemption. The use of VA data was reviewed by the Department of Veterans Affairs Central IRB, was determined to meet the criteria for exemption under Exemption Category 4(3), and approved for Waiver of HIPAA Authorization. The use of SIDIAP was approved by the Clinical Research Ethics Committee of the IDIAPJGol (project code: 20/070-PCV). The use of CPRD was approved by the Independent Scientific Advisory Committee (ISAC) (protocol number 20_059RA2). The use of IQVIA-OpenClaims was exempted from IRB approval.

## Data Sharing Statement

Analyses were performed locally in compliance with all applicable data privacy laws. Although the underlying data is not readily available to be shared, authors contributing to this paper have direct access to the data sources used in this study. All results (e.g. aggregate statistics, not presented at a patient-level with redactions for minimum cell count) are available for public inquiry. These results are inclusive of site-identifiers by contributing data sources to enable interrogation of each contributing site. All analytic code and result sets are made available at: https://github.com/ohdsistudies/Covid19CharacterizationCharybdis

## Notes

□ Conflict of Interest: Author reports a potential financial conflict of interest associated with the Observational Health Data Science & Informatics Characterizing Health Associated Risks and Your Baseline Disease In SARS-COV-2 (CHARYBDIS) study group. Specifically, author received grants from Innovative Medicines Initiative and Janssen Research and Development in support of this work.

### Competing Interest Statement

DPA, PJR, PBR, AS report a potential financial conflict of interest associated with the Observational Health Data Science & Informatics Characterizing Health Associated Risks and Your Baseline Disease In SARS-COV-2 (CHARYBDIS) study group. Specifically, author received grants from Innovative Medicines Initiative and Janssen Research and Development in support of this work. Unless otherwise indicated, authors certify that they have NO affiliations with or involvement in any organization or entity with any financial interest or non-financial interest in the subject matter or materials discussed in this manuscript.

### Clinical Protocols

https://github.com/ohdsi-studies/Covid19CharacterizationCharybdis/blob/master/documents/Protocol_COVID-19%20Charybdis%20Characterisation_V5.docx

### Funding Statement

The European Health Data & Evidence Network has received funding from the Innovative Medicines Initiative 2 Joint Undertaking (JU) under grant agreement No 806968. The JU receives support from the European Unions Horizon 2020 research and innovation programme and EFPIA. This research received partial support from the National Institute for Health Research (NIHR) Oxford Biomedical Research Centre (BRC), US National Institutes of Health, US Department of Veterans Affairs, Janssen Research & Development, and IQVIA. The University of Oxford received funding related to this work from the Bill & Melinda Gates Foundation (Investment ID
INV-016201 and INV-019257). VINCI [VA HSR RES 13-457] (SLD, MEM, KEL). JCEL has received funding from the Medical Research Council (MR/K501256/1) and Versus Arthritis (21605). No funders had a direct
role in this study. The views and opinions expressed are those of the authors and do not necessarily
reflect those of the Clinician Scientist Award programme, NIHR, Department of Veterans Affairs or
the United States Government, NHS, or the Department of Health, England. 

